# Early-Life Sugar Restriction, Multi-omics Architecture, and Multisystem Resilience: A Natural Experiment

**DOI:** 10.64898/2026.04.30.26352133

**Authors:** Yiwei Zhang, Dan Chen, Xiaolong Liang, Xianglian Cai, Ziliang Ye, Yanjun Zhang, Sisi Yang, Xiaoqin Gan, Yu Huang, Yiting Wu, Yuanyuan Zhang, Xianhui Qin

## Abstract

**Background:** The systemic long-term health effects and underlying biological mechanisms of early-life sugar restriction remain poorly defined.

**Methods:** We exploited a natural experiment created by the abrupt end of UK postwar sugar rationing (September 1953), including 60,768 UK Biobank participants born between October 1951 and March 1956. Exposure to early-life sugar restriction was defined by birth date relative to the policy end. We assessed 27 incident disease outcomes and all-cause mortality. In subsets, we performed plasma proteomic and metabolomic profiling, evaluated 81 adult phenotypes, and applied formal mediation analysis.

**Findings:** Of 60,768 participants (mean age 54.6 years; 56.2% female), 38,453 (63.3%) were exposed to early-life sugar restriction. Longer exposure was associated with dose-dependent risk reductions for infections, cancer, mental and behavioural disorders, nervous system, digestive, musculoskeletal, genitourinary, and skin disorders (adjusted HRs 0.83–0.93), and with lower all-cause mortality (adjusted HR 0.79; 95% CI 0.73–0.87). The exposure was associated with a distinct molecular signature and an adult phenotype marked by higher fat-free mass and lower basal metabolic rate, with no difference in BMI. Mediation analyses identified a modest “molecular memory” pathway (3–9% of effect) and a dominant “physiological programming” pathway (4–13% of effect).

**Interpretation:** Sugar restriction in the first 1,000 days programs multisystem resilience that substantially reduces risk of chronic diseases and later-life mortality. This protection operates through a hierarchical biological architecture dominated by a metabolically efficient physiological phenotype, providing mechanistic support for stricter regulation of added sugars in infant foods.

**Fundings:** National Natural Science Foundation of China and other funding sources.

**Research in Context:** *Evidence before this study:* We searched PubMed, Web of Science, and medRxiv up to January 31, 2026, using “sugar rationing”, “natural experiment”, and “UK Biobank”. Studies using the 1953 end of UK sugar rationing as a natural experiment have reported reduced risks of type 2 diabetes, hypertension, cardiovascular disease, metabolic dysfunction-associated steatotic liver disease, respiratory conditions, heart failure, and anxiety. All previous studies examined single disease outcomes in isolation; none tested for coordinated protection across organ systems or examined underlying biological mechanisms with multi-omics profiling.

*Added value of this study:* This study demonstrates that early-life sugar restriction confers dose-dependent protection across 27 incident outcomes spanning multiple organ systems—including infections, cancer, mental and behavioural disorders, nervous system, digestive, musculoskeletal, and genitourinary diseases—and reduces all-cause mortality, establishing a pattern of multisystem resilience. Through proteomic and metabolomic profiling, it identifies a molecular signature of early-life sugar restriction and a corresponding adult physiological phenotype characterized by higher fat-free mass and lower basal metabolic rate, with no difference in BMI. Formal mediation analysis reveals a hierarchical dual-pathway mechanism: a modest molecular memory (3–9% of total effect) and a dominant physiological programming pathway mediated by fat-free mass and basal metabolic rate (4–13%). Conventional risk traits, including visceral adiposity and dysglycemia, showed no significant mediation.

*Implications of all the available evidence:* Early-life nutrition programs lifelong multisystem resilience through a hierarchical biological architecture dominated by physiological reprogramming. These findings provide a mechanistic mandate for stricter regulation of added sugars in infant and toddler foods as primary prevention of non-communicable diseases, and highlight fat-free mass and basal metabolic rate as physiological pathways for future risk assessment and intervention.

## Introduction

The first 1,000 days after conception are a critical window for developmental programming, where nutritional exposures durably shape physiology [1,2]. The UK’s postwar sugar rationing, which ended abruptly in September 1953, provides a rare natural experiment. During rationing, sugar intake was restricted to levels aligning with today’s dietary guidelines [3,4]; afterwards, consumption nearly doubled. Because the policy’s end created a sharp, quasi-random discontinuity in early-life sugar exposure based solely on birth date, it has been increasingly used to infer causal health effects [5,6].

However, two critical gaps remain. First, whether this protection extends beyond cardiometabolic disease is unknown. All existing studies have examined single outcomes—spanning type 2 diabetes, hypertension [5], cardiovascular disease [6], liver disease [7], respiratory health [8], heart failure [9], and mental health [10]—but no study has tested whether early-life sugar restriction concurrently reduces risk across multiple organ systems. If early nutrition calibrates core systemic physiology, protection should manifest as multisystem resilience across neuropsychiatric, neoplastic, immune, and musculoskeletal domains. Second, the mediating biological pathways are unknown. Two hypotheses exist: “molecular memory” (durable shifts in the adult proteome and metabolome) [11] and “physiological programming” (permanent recalibration of body composition and energy metabolism), but their relative contributions have never been empirically quantified.

To resolve these questions, we applied the UK sugar rationing natural experiment to the UK Biobank, combining prospective disease phenomics across 27 outcomes, plasma proteomics (2,923 proteins), metabolomics (168 metabolites), and comprehensive adult phenotyping in 60,768 participants. We aimed to: (1) map whether early-life sugar restriction confers multisystem resilience beyond cardiometabolic endpoints; (2) identify associated molecular signatures in the adult proteome and metabolome; and (3) quantify, through formal causal mediation analysis, the relative contributions of molecular memory versus physiological programming pathways. By simultaneously testing breadth and mechanism, this design moves beyond single-disease associations to define the biological architecture of programmed resilience.

## Methods

### Study Design and Population

We exploited a natural experiment using the abrupt nationwide termination of the UK’s sugar rationing policy on 30 September 1953, which created a sharp, quasi-random discontinuity in early-life dietary sugar exposure based solely on birth date. We defined the first 1,000 days from conception as the exposure window. Using an average quarter length of 91 days, this resulted in an exposure period from 1 October 1951 to 30 June 1954. Individuals born on or after 1 July 1954 were classified as unexposed [5,6] (**Figure 1A**).

**Figure 1.**
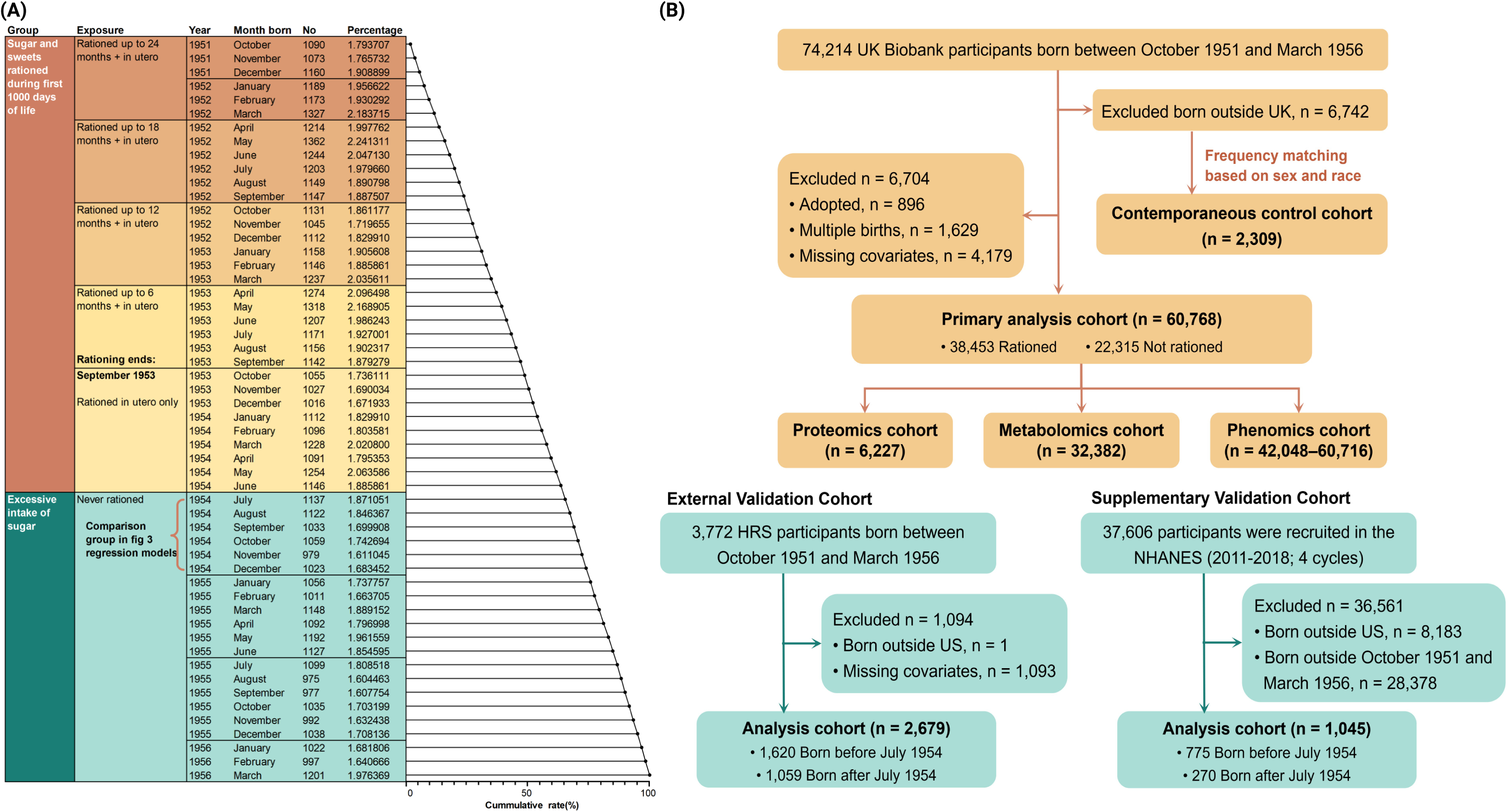
Sample Distribution by Birth Month and Prenatal Sugar Rationing Exposure and Study Cohort Flowchart. (**A**) Distribution of births by calendar month and exposure to prenatal sugar rationing. Orange bars represent participants exposed to prenatal sugar rationing during pregnancy. Green bars represent unexposed participants, who served as the control group for assessing associations between early-life nutritional restriction and multisystem health outcomes. (**B**) Flowchart of the study cohort. **Abbreviations:** HRS, Health and Retirement Study; NHANES, National Health and Nutrition Examination Survey.

From the UK Biobank, we identified eligible participants born between October 1951 and March 1956 (n = 74,214). After excluding individuals born outside the UK, adopted, part of a multiple birth, or with missing key covariates, the primary analytical cohort comprised 60,768 participants. Within this cohort, 38,453 were classified as exposed to early-life sugar restriction and 22,315 as unexposed (**Figure 1B**). Subsets with proteomic (n = 6,227), metabolomic (n = 32,382), and deep phenomic (n = 42,048–60,716) data were used for mechanistic analyses.

### Exposure Definition and Outcome Assessment

To model a biological dose-response, exposure was categorized into four mutually exclusive groups based on the duration of exposure within the first 1000 days: (1) never exposed; (2) exposed *in utero* only; (3) exposed *in utero* and up to one year postnatally; (4) exposed *in utero* and for one to two years postnatally.

We prospectively assessed 27 incident clinical endpoints, including 11 broad disease categories, 11 specific diseases, all-cause mortality, and four cause-specific mortalities [12]. Outcomes were defined using ICD-10 codes and ascertained from linked national electronic health records (primary care, hospital inpatient, death registries).

### Covariate Adjustment and Validation Cohorts

Covariates were adjusted for in two sequential models. Model 1 included sex and race. Model 2 additionally adjusted for place of birth, deciles of birth coordinates, birth month, parental history of diabetes and cardiovascular disease, and year of baseline assessment. For relevant disease outcomes, Model 2 also included corresponding polygenic risk scores [13].

To test the specificity of associations, we employed multiple validation strategies. An internal negative control cohort comprised UK Biobank participants born outside the UK (n=2,309). We also utilized two external US cohorts, classifying individuals into an analogous birth-era exposure based on the same temporal cutoff (1 July 1954): the Health and Retirement Study (HRS; n=2,679, lnorm models) and the National Health and Nutrition Examination Survey (NHANES; n=1,045, logistic regression for prevalent conditions) (See **Supplementary Methods** for full cohort specifications.)

### Statistical Analysis

Associations between rationing exposure and disease risk were evaluated using Gompertz parametric survival models, which provided the best fit (lowest Akaike Information Criterion) for most endpoints. To elucidate potential biological pathways, we implemented a multi-step mediation pipeline: (1) LASSO regression selected exposure-associated molecular features; (2) linear models identified exposure-associated phenotypic traits (FDR-corrected); (3) candidates were screened for disease association; and (4) formal causal mediation analysis quantified the proportion of the total effect attributable to each mediator. Sensitivity analyses tested the robustness of the primary associations to further covariate adjustment.

For complete technical specifications regarding model selection, validation cohorts, phenotypic and multi-omics profiling, and bioinformatics analyses, refer to the **Supplementary Methods**.

## Results

### Study Population and Baseline Characteristics

The primary analysis included 60,768 UK Biobank participants (mean age 54.6 ± 1.6 years; 56.2% female; 99.3% White) born between October 1951 and March 1956. Of these, 38,453 (63.3%) were classified as exposed to early-life sugar rationing and 22,315 (36.7%) as unexposed (**Table 1**).

**Table 1.** Baseline characteristics of the study population by early-life exposure to sugar rationing. *.

| Characteristics | Overall<br>(n = 60,768) | Rationed<br>(n = 38,453) | Not rationed<br>(n = 22,315) | *P* value |
| --- | --- | --- | --- | --- |
| Age assesses, years | 54.6 (1.6) | 55.4 (1.2) | 53.2 (1.0) | <0.001 |
| Female, n (%) | 34136 (56.2) | 21552 (56.0) | 12584 (56.4) | 0.414 |
| White, n (%) | 60333 (99.3) | 38217 (99.4) | 22116 (99.1) | <0.001 |
| College/university, n (%) | 21573 (35.5) | 13563 (35.3) | 8010 (35.9) | <0.001 |
| Household (52,000-100,000), n (%) | 14859 (27.5) | 9025 (26.5) | 5834 (29.2) | <0.001 |
| Townsend deprivation index | -1.5 (3.0) | -1.5 (3.0) | -1.4 (3.0) | <0.001 |
| Place of birth, n (%) |  |  |  | 0.002 |
| England | 52089 (85.7) | 33061 (86.0) | 19028 (85.3) |  |
| Scotland | 5661 (9.3) | 3462 (9.0) | 2199 (9.9) |  |
| Wales | 3018 (5.0) | 1930 (5.0) | 1088 (4.9) |  |
| Birth season, n (%) |  |  |  | <0.001 |
| Spring | 15938 (26.2) | 11305 (29.4) | 4633 (20.8) |  |
| Summer | 13736 (22.6) | 8276 (21.5) | 5460 (24.5) |  |
| Autumn | 14785 (24.3) | 8710 (22.7) | 6075 (27.2) |  |
| Winter | 16309 (26.8) | 10162 (26.4) | 6147 (27.5) |  |
| Birth weight, kg | 3.3 (0.6) | 3.3 (0.6) | 3.3 (0.6) | 0.703 |
| Breastfed, n (%) | 35708 (73.3) | 22676 (74.0) | 13032 (72.0) | <0.001 |
| Maternal smoking, n (%) | 18713 (35.1) | 11838 (35.2) | 6875 (35.0) | 0.556 |
| Physical activity, METs/week | 2520 (2701) | 2516 (2682) | 2527 (2734) | 0.638 |
| Smoking status, n (%) |  |  |  | <0.001 |
| Never | 34167 (56.4) | 21444 (55.9) | 12723 (57.2) |  |
| Previous | 19680 (32.5) | 12779 (33.3) | 6901 (31.0) |  |
| Current | 6754 (11.1) | 4117 (10.7) | 2637 (11.8) |  |
| Alcohol consumption status, n (%) |  |  |  | <0.001 |
| <1 time/week | 16313 (26.9) | 10361 (27.0) | 5952 (26.7) |  |
| 1-2 times/week | 16438 (27.1) | 10271 (26.7) | 6167 (27.7) |  |
| 3-4 times/week | 15511 (25.5) | 9695 (25.2) | 5816 (26.1) |  |
| >4 times/week | 12465 (20.5) | 8104 (21.1) | 4361 (19.6) |  |
| Body mass index, kg/m<sup>2</sup> | 27.5 (5.0) | 27.6 (5.0) | 27.5 (5.0) | 0.137 |
| Waist circumference, cm | 90.1 (13.9) | 90.3 (13.8) | 89.9 (13.9) | <0.001 |
| Healthy diet score | 3.0 (1.4) | 3.0 (1.4) | 3.0 (1.4) | <0.001 |
| Healthy sleep score | 3.0 (1.0) | 3.0 (1.0) | 3.0 (1.0) | 0.021 |
| Healthy mental score | 4.6 (3.3) | 4.6 (3.2) | 4.6 (3.3) | 0.003 |
| HbA1c, mmol/mol | 35.8 (6.4) | 36.0 (6.5) | 35.5 (6.2) | <0.001 |
| Systolic blood pressure, mmHg | 136.3 (17.6) | 136.8 (17.6) | 135.3 (17.4) | <0.001 |
| Triglycerides, mmol/L | 1.8 (1.0) | 1.8 (1.0) | 1.7 (1.1) | 0.002 |
| Cholesterol, mmol/L | 5.9 (1.1) | 5.9 (1.1) | 5.8 (1.1) | <0.001 |
| **HDL cholesterol, mmol/L** | 1.5 (0.4) | 1.5 (0.4) | 1.5 (0.4) | 0.371 |
| **LDL cholesterol, mmol/L** | 3.7 (0.9) | 3.7 (0.9) | 3.7 (0.8) | 0.002 |
| **C-reactive protein, mg/L** | 2.5 (4.2) | 2.5 (4.3) | 2.5 (4.0) | 0.03 |
| **Parental history of cardiovascular disease, n (%)** | 45024 (74.1) | 28478 (74.1) | 16546 (74.1) | 0.818 |
| **Parental history of diabetes, n (%)** | 11283 (18.6) | 6993 (18.2) | 4290 (19.2) | 0.002 |

\*Variables are presented as mean (SD) or n (%).

Compared with the unexposed group, the rationed cohort was slightly older (55.4 *vs.* 53.2 years) and had a higher proportion of summer births. This group also had marginally lower household income, lower Townsend deprivation index scores, and a lower prevalence of parental history of diabetes. Sex distribution, race, and birth weight were balanced between groups. In adulthood, the rationed group exhibited subtly higher levels of HbA1c, systolic blood pressure, and triglycerides. The demographic and clinical profiles of the proteomic and metabolomic sub-samples were consistent with the primary cohort (**Table S5**).

### Association of Early-Life Sugar Rationing Exposure with Multisystem Disease Risk

#### Primary Dose-Response Associations

**Table 2** and **Table S6** summarize associations between early-life sugar rationing exposure and disease risk across multiple organ systems. After adjustment for confounders (Model 2), a longer rationing duration was associated with progressively lower risks for most endpoints, with statistically significant trend tests (*P* for trend <0.05).

**Table 2.**
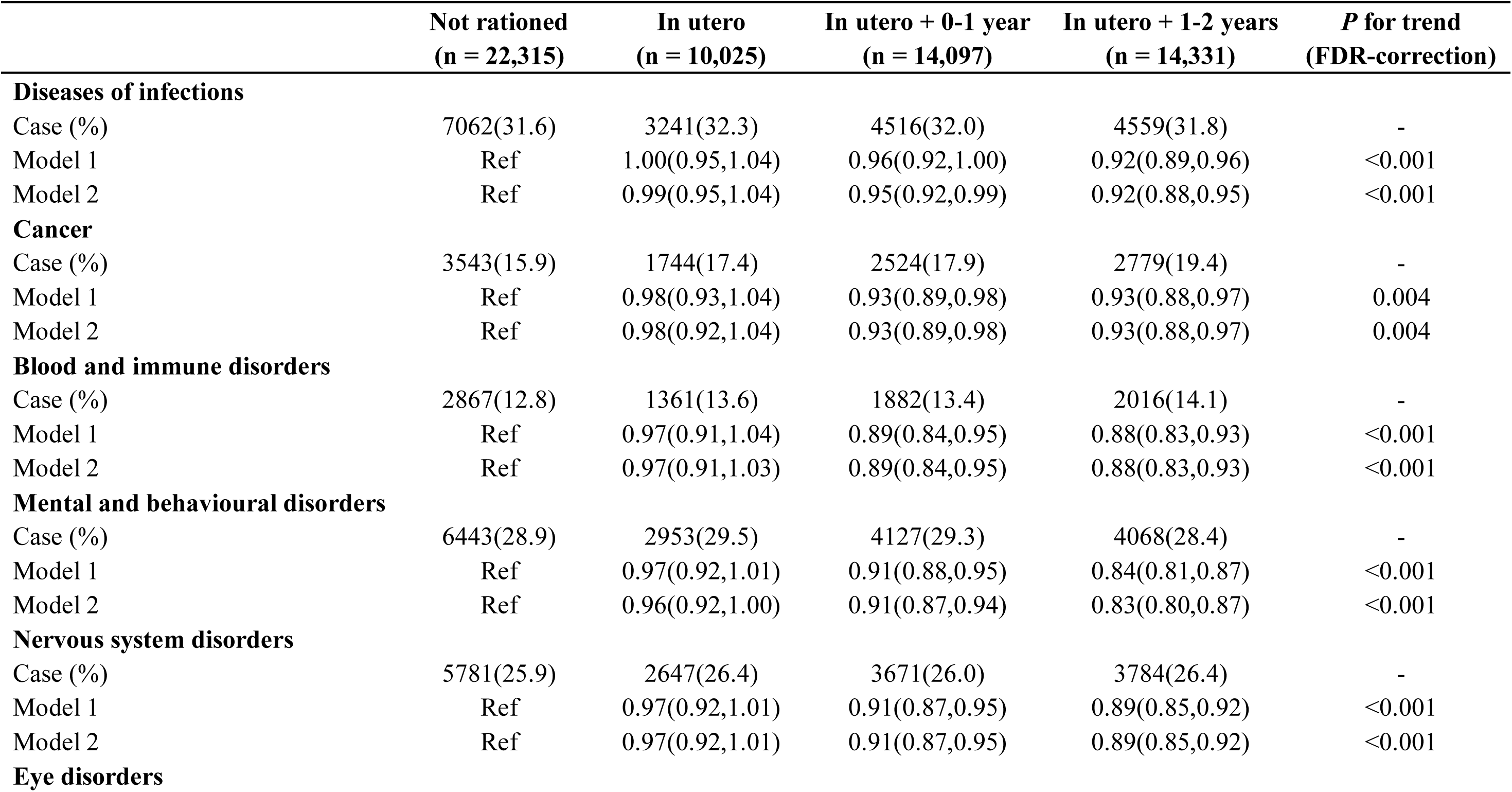
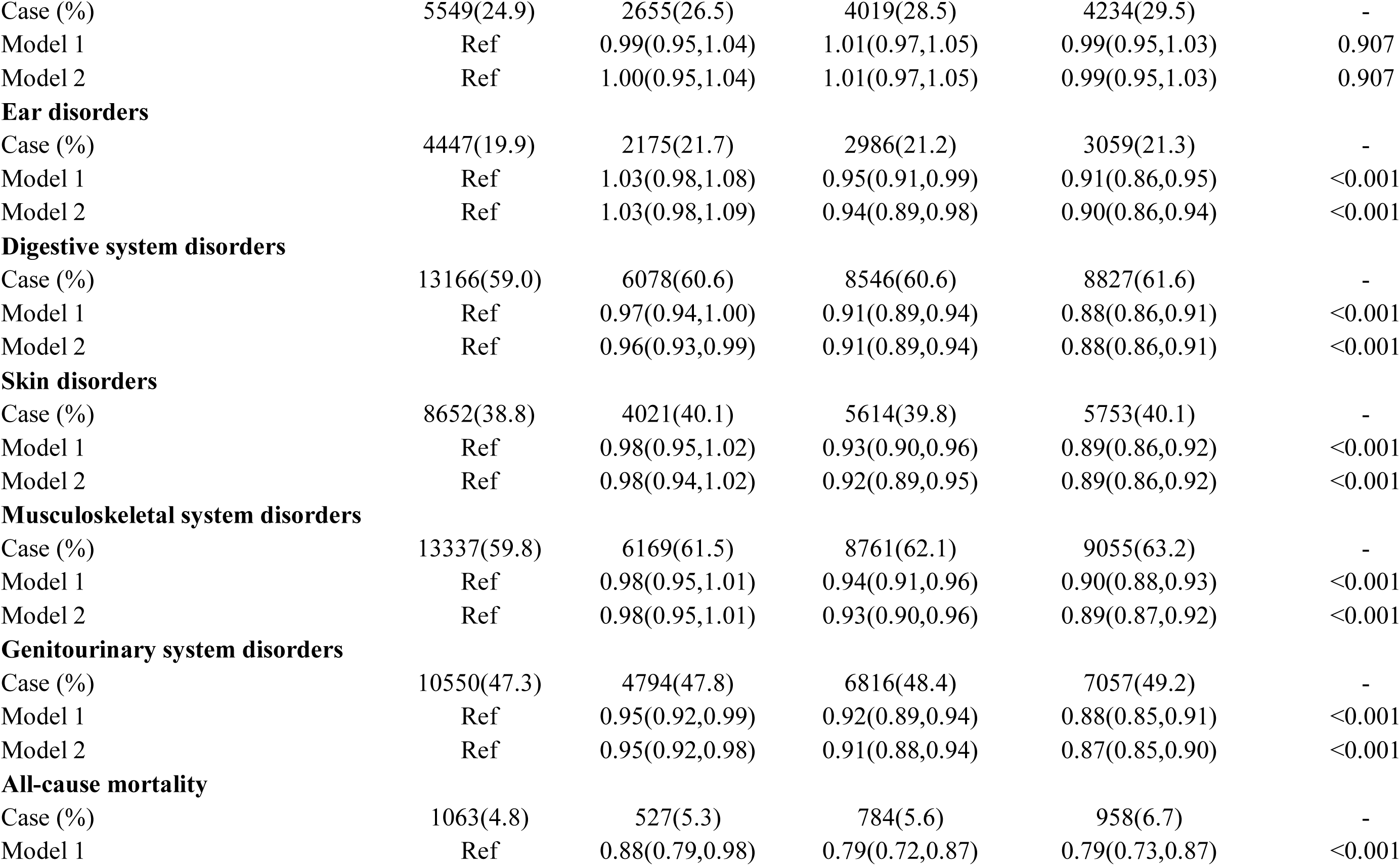
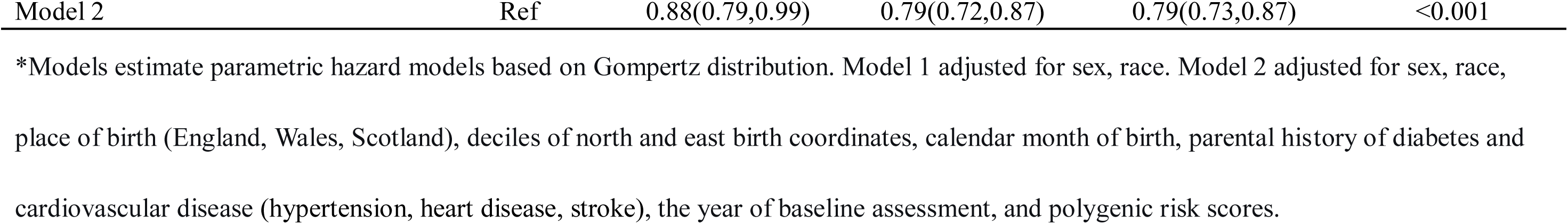
Associations of early-life sugar restriction with incident multisystem diseases (broad disease categories). *.

Compared to the unexposed group, individuals with the longest exposure (in utero plus 1-2 years) had significantly reduced risks for numerous major disease categories, including infections (adjusted, HR 0.92; 95%CI, 0.88-0.95), cancer (0.93, 0.88-0.97), blood and immune disorders (0.88, 0.83-0.93), mental and behavioral disorders (0.83, 0.80-0.87), nervous system disorders (0.89, 0.85-0.92), ear disorders (0.90, 0.86-0.94), digestive disorders (0.88, 0.86-0.91), skin disorders (0.89, 0.86-0.92), musculoskeletal disorders (0.89, 0.87-0.92), and genitourinary disorders (0.87, 0.85-0.90) (**Table 2**). Similar inverse trends were observed for specific conditions such as bacterial/viral infections, anemia, dementia, mood/neurotic disorders, sleep disorders, inflammatory bowel disease, liver diseases, and renal failure (adjusted HR range 0.78-0.93) (**Table S6)**. Early-life sugar rationing was also associated with lower all-cause mortality (0.79, 0.73-0.87) and cause-specific mortality from cancer (0.85, 0.76-0.96) and circulatory diseases (0.78, 0.66-0.91). In contrast, no significant association was observed for eye disorders (*P* for trend = 0.907) (**Table 2**).

#### Temporal Dynamics via Event Study

An event-study design tracking risk across finer temporal windows relative to the end of rationing corroborated the dose-response pattern (**Figure 2)**. Using individuals born in the 9 months immediately before rationing ended as the reference (HR = 1), adjusted HRs declined progressively with longer postnatal exposure for nearly all disease systems. Significant downward trends emerged after 6–12 months of postnatal exposure and persisted through the longest exposure window (in utero plus 24 months). This pattern was evident for mental/behavioral, nervous, digestive, skin, musculoskeletal, genitourinary, and ear disorders, as well as cancer. In contrast, the adjusted HRs for eye disorders remained close to unity across all periods. All-cause mortality exhibited the steepest graded reduction.

**Figure 2.**
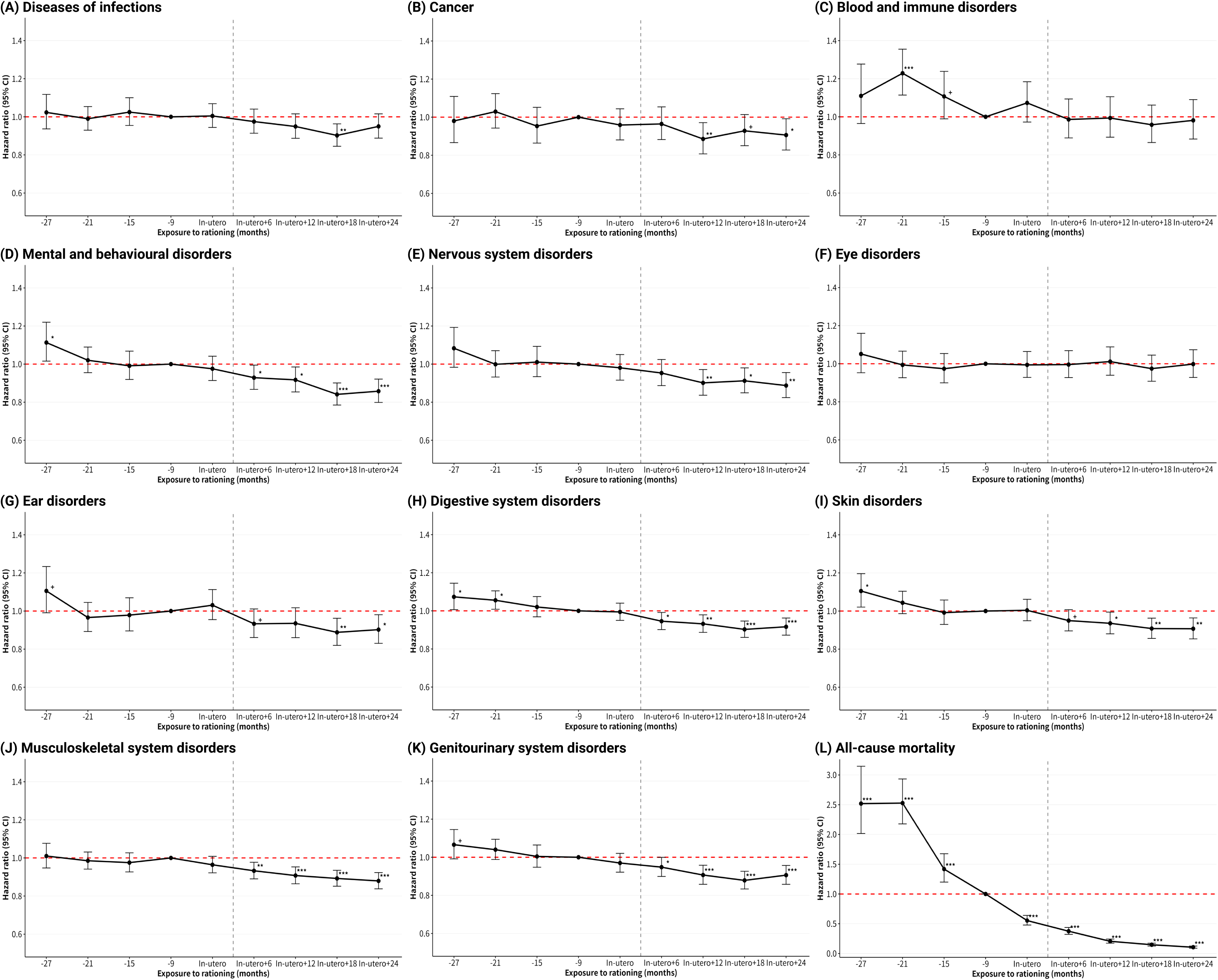
Hazard Ratios of Health Outcomes According to Timing and Duration of Prenatal and Early-Life Sugar Rationing Exposure. Each point shows the hazard ratio comparing adults exposed to sugar rationing in utero, in utero plus 6, 12, 18, or 24 months after birth, or never exposed, to the reference group (born July–December 1954, >9 months after rationing ended). Never-exposed adults also include those born January–June 1955 (>15 months after rationing), July–December 1955 (>21 months after rationing), and January–March 1956 (>27 months after rationing). All models are adjusted for sex, race, place of birth (England, Wales, Scotland), deciles of north and east birth coordinates, calendar month of birth, parental history of diabetes and cardiovascular disease (hypertension, heart disease, stroke), year of baseline assessment, and polygenic risk scores. Asterisks denote statistical significance: +*P* < 0.1, \**P* < 0.05, \*\**P* < 0.01, \*\*\**P* < 0.001.

#### Validation and Sensitivity Analyses

Validation cohorts revealed no spurious associations. In the internal negative control cohort of non-UK-born individuals (inherently unexposed), no association was found between birth-era classification and disease risk (**Tables S7**). Similarly, in the external US cohorts (HRS and NHANES), where individuals were classified into “born before July 1954” or “born after July 1954” based solely on birth date, we observed no consistent protective pattern akin to that in the primary UK analysis (**Tables S8-S9**).

The main associations were robust to further adjustment for early-life factors (breastfeeding, maternal smoking, birth weight) and adult socioeconomic/lifestyle factors (**Table S10**). Stratified analyses demonstrated generally consistent effects across subgroups defined by sex, race, place/season of birth, and polygenic risk scores (**Figure S1**). Isolated significant interactions for sex (blood and immune disorders and genitourinary disorders) were likely due to chance given multiple testing.

### Omics Signatures Associated with Early-Life Sugar Restriction

#### Molecular Signatures

LASSO regression identified 104 plasma proteins and 31 metabolites significantly associated with early-life sugar restriction (**Tables S11-S12**). Enrichment analysis of these proteins revealed overrepresentation in 61 pathways, including extracellular region, extracellular space, extracellular matrix, hormone activity, extracellular matrix structural constituent, and positive regulation of MAPK cascade (**Figure 3A, Table S13**). Metabolites were enriched in 24 metabolic pathways, most notably glycine/serine/threonine metabolism, one carbon pool by folate, glycine/serine metabolism, and phenylalanine/tyrosine/tryptophan biosynthesis (**Figure 3B, Table S14**). PPI network analysis identified fibroblast growth factor 7 (FGF7) as the central hub, exhibiting the highest degree and betweenness centrality (**Figure 3C, Table S15**).

**Figure 3.**
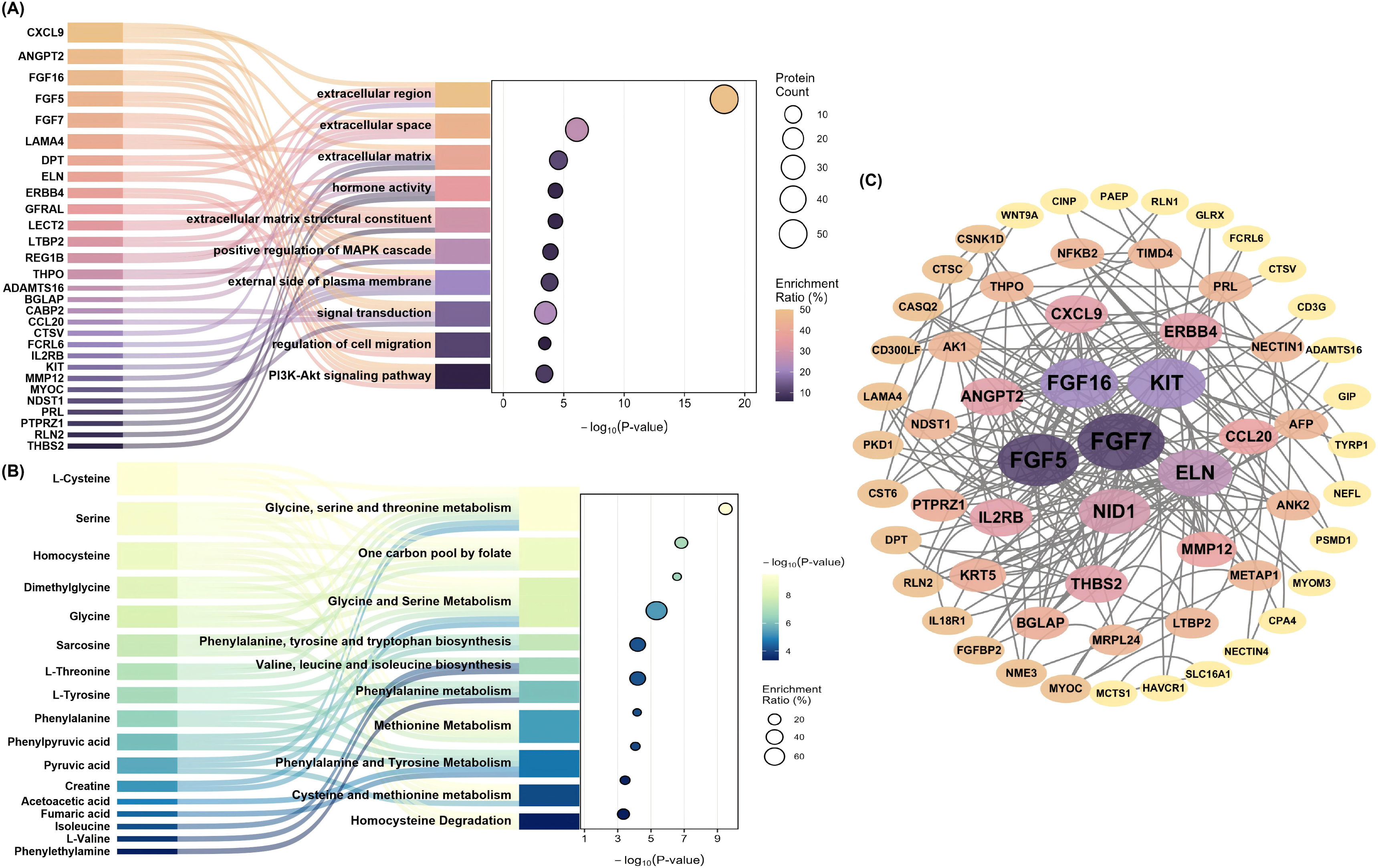
Functional, Pathway, and Protein-Protein Interaction (PPI) Network Analyses of Early-Life Sugar Restriction Signatures. **(A)** Functional and Pathway Enrichment Analysis of Proteomic Signatures.Top ten significantly enriched pathways are shown, with five representative proteins per pathway. **(B)** Functional and Pathway Enrichment Analysis of Metabolomic Signatures. Top ten significantly enriched pathways are shown, with representative metabolites per pathway. **(C)** Protein-Protein Interaction (PPI) Network Analysis of Proteomic Signatures.

#### Adult Phenotypic Associations

A comprehensive screen of 81 quantitative adult phenotypes identified 50 traits that remained significantly associated with early-life sugar restriction after FDR correction (**Figure S2, Table S16**). The association profile was characterized by widespread, subtle shifts across physiological domains, with the most pronounced statistical signals (all *P* < 10⁻³⁰) indicating alterations in respiratory and renal function: modestly lower lung function (FEV₁ [ forced expiratory volume in 1-second]: β = −0.033; FVC [forced vital capacity]: β = −0.036) and elevated cystatin C (β = 0.007), a marker of reduced glomerular filtration rate. These early-life exposure-related alterations in distinct organ systems highlight the multisystem nature of the programming effect.

Notably, body mass index (BMI) showed no significant association, suggesting that the long-term health effects are not simply mediated by differences in general adiposity. Instead, the exposure was linked to a distinct pattern encompassing measures of central adiposity (e.g., waist circumference), lower basal metabolic rate, higher fat-free mass, and specific differences in circulating lipids, liver enzymes, and inflammatory indices (**Table S16**).

### Association between Omics Signatures and Multisystem Diseases

#### Molecular Associations with Disease Risk

Candidate proteins and metabolites identified as exposure-associated were tested for links to incident disease. Several exhibited significant, directionally consistent associations across multiple outcomes (**Figure S3, Tables S17-S18**). For example, proteins Hepatitis A virus cellular receptor 1 (HAVCR1), Angiopoietin-2 (ANGPT2), and CUB domain-containing protein 1 (CDCP1), along with metabolites Glycoprotein Acetyls (GlycA) and glucose, were positively associated with risk of mental/behavioral disorders and all-cause mortality. Conversely, carbonic anhydrase 14 (CA14) and metabolites such as total cholesterol showed inverse associations.

#### Phenotypic Associations with Disease Risk

Similarly, exposure-associated phenotypic traits were examined for disease relationships (**Figure S3, Table S19**). Measures of visceral adiposity (e.g., visceral adiposity index, waist circumference), glucose metabolism (e.g., HbA1c, TyG index), systemic inflammation (e.g., systemic immune-inflammation index, platelet-to-lymphocyte ratio), and renal function (cystatin C) were robustly and positively associated with higher risk across a broad spectrum of conditions, including infectious diseases and cancer.

### Mediation Pathways of Protection

For the “molecular memory” pathway, 62 proteins and 31 metabolites met criteria for mediation analysis; for the “physiological programming” pathway, 50 phenotypic traits were tested. Causal mediation analyses identified significant indirect effects in both pathways.

Molecular mediators included plasma proteins that mediated 5-8% of the protection for nervous and blood/immune disorders—notably glycodelin (PAEP), cytokine receptor-like factor 1 (CRLF1), and receptor-type tyrosine-protein phosphatase zeta (PTPRZ1) (**Figure 4, Table S20**). Key metabolites implicated in mitochondrial function (citrate) and lipid metabolism (fatty acid unsaturation, omega-3 fatty acids) mediated 3-9% of effects for mental, nervous, and digestive disorders (**Figure 4, Table S21**).

**Figure 4.**
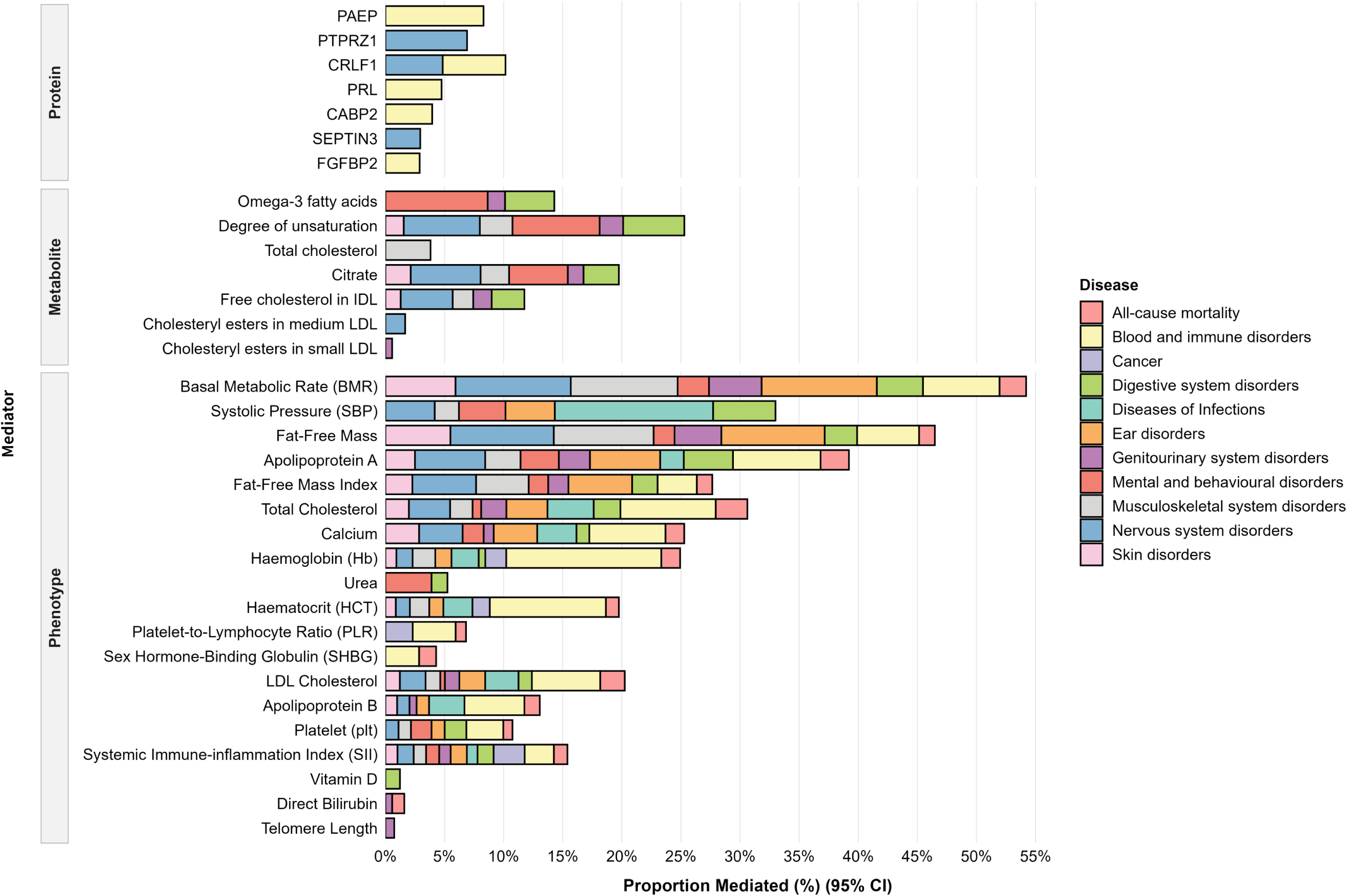
Mediation Analysis of Multi-omic Signatures in the Association Between Early-Life Sugar Restriction and Multisystem Diseases. All regression models are adjusted for sex, race, place of birth (England, Wales, Scotland), deciles of north and east birth coordinates, calendar month of birth, parental history of diabetes and cardiovascular disease (hypertension, heart disease, stroke), year of baseline assessment, and polygenic risk scores.

Phenotypic mediators accounted for larger proportions of the protective associations. Basal metabolic rate (BMR) and fat-free mass were the most prominent, mediating 4-13% of the effect across nervous, musculoskeletal, skin, and genitourinary disorders. Systolic blood pressure (SBP) mediated 4-13% of the effect across infections, mental, nervous, and digestive disorders. Specific circulating lipids (e.g., apolipoprotein A) and inflammatory indices (e.g., platelet-to-lymphocyte ratio) also mediated effects for infections, cancer, ear disorders, and mortality. In contrast, conventional risk traits such as visceral adiposity and dysglycemia showed no significant mediation (**Figure 4, Table S22**).

## Discussion

In this natural experiment, longer exposure to sugar restriction in early life was associated with progressively lower risks of incident disease across multiple organ systems and with reduced all-cause mortality. This broad, dose-dependent protection points to a programmed state of multisystem resilience rather than isolated disease-specific effects. We further identified two complementary biological pathways underlying this protection: a modest molecular memory and a dominant physiological reprogramming involving body composition and energy metabolism. These findings raise several interconnected issues for discussion.

### A Paradigm of Multisystem Resilience

Our primary finding is the breadth of protection conferred by early-life sugar restriction. The consistent, dose-dependent risk reductions spanned mental, nervous, digestive, and musculoskeletal disorders—extending far beyond the single-disease outcomes examined in previous studies using this natural experiment [5–10]. This breadth argues against a disease-specific mechanism and instead points to a shared biological substrate, which we term multisystem resilience.

The null association with eye disorders reinforces this interpretation. Eye disorders share few developmental pathways with the conditions that showed protection, serving as a disease-domain negative control that rules out a non-specific, all-organ effect. The temporal dynamics are similarly informative: protective trends emerged only after 6–12 months of postnatal exposure, coinciding with the introduction of solid foods [14]. This timing pinpoints a postnatal window in which nutritional programming actively shapes long-term resilience, independent of in-utero exposure alone.

### The Biological Imprint of Early Nutrition

Our multi-omic analyses identified a molecular signature associated with early-life sugar restriction. The proteomic profile was enriched for pathways governing tissue communication and extracellular matrix organization, with FGF7 emerging as a central network hub [15,16]. The metabolomic signature indicated shifts in amino acid metabolism and glycolytic pathways, suggesting a lasting recalibration of core bioenergetics [17,18].

These molecular features were mirrored in a distinct adult physiological phenotype. Body mass index showed no association with the exposure. Instead, early-life sugar restriction was linked to higher fat-free mass, lower basal metabolic rate, and subtle shifts in organ system function, including lung function and renal filtration markers [19,20]. Together, this molecular and physiological imprint constitutes the biological substrate of the observed resilience. The critical question is through which pathway this protection is primarily mediated.

### Relative Contributions of Molecular Memory and Physiological Programming

Our mediation analysis quantified the contributions of the two hypothesized pathways. The ‘physiological programming’ pathway was dominant: basal metabolic rate and fat-free mass mediated 4–13% of the protective effect across nervous, musculoskeletal, skin, and genitourinary disorders. By contrast, the “molecular memory” pathway contributed modestly, with specific plasma proteins (e.g., CRLF1, PTPRZ1) and metabolites (e.g., citrate, fatty acid unsaturation) mediating 5–8% and 3–9% of the effect for blood, immune, and nervous system disorders, respectively. Visceral adiposity and dysglycemia showed no significant mediation, dissociating the principal programming effect from conventional cardiometabolic risk factors.

These findings suggest a hierarchical biological architecture: a metabolically efficient physiological phenotype—higher fat-free mass, lower basal metabolic rate—constitutes the dominant route of protection, while durable molecular imprints provide a modest, persistent background signal. This quantitative hierarchy may inform the development of interventions that target physiological programming rather than molecular memory alone.

### Clinical and Public Health Implications

These findings suggest that the goal of early-life nutrition should extend beyond supporting growth to programming lifelong, multisystem resilience. Limiting added sugar exposure in the first 1,000 days may reduce risk for a broad spectrum of conditions—extending well beyond the single-disease endpoints examined in previous studies [5–10]—including mental health disorders, dementia, and cancer, which collectively dominate later-life morbidity and mortality.

For clinical practice, the identified mediators—particularly the dominant role of fat-free mass and basal metabolic rate—highlight physiological pathways that could inform future risk assessment and intervention. For public health, this study strengthens the evidence base for policies that restrict added sugars in commercial infant and toddler foods. The dose-dependent protection observed across organ systems suggests that such measures represent a population-wide investment in the primary prevention of non-communicable disease.

### Limitations

Several limitations warrant consideration. First, although the regression discontinuity design controls for smoothly varying confounders, unmeasured factors that changed abruptly at the policy cutoff cannot be ruled out; historical analyses, however, identify no major concurrent shifts. Second, individual-level dietary data are unavailable, and sugar consumption rose sharply after rationing ended, exceeding pre-war levels. This implies that the unexposed reference group likely experienced unusually high early-life sugar intake, making our effect estimates conservative. Third, if sugar restriction reduced childhood mortality, selective survival of frailer individuals into the exposed cohort would similarly bias associations toward the null. The robust dose-dependent protection we observe despite these biases supports the strength of the underlying biological programming. Fourth, proteomic, metabolomic, and phenotypic data were measured cross-sectionally at mid-life, precluding analysis of life-course trajectories and limiting causal inference regarding their role as mediators. Upstream developmental biology linking early sugar exposure to the identified mediators requires functional validation in model systems.

### Conclusions

In this natural experiment, sugar restriction during the first 1,000 days was associated with substantial, dose-dependent reductions in incident disease across multiple organ systems and with lower all-cause mortality. This protection operated not through conventional cardiometabolic risk factors but through a hierarchical dual-pathway mechanism: a modest molecular memory in the plasma proteome and metabolome, and a dominant physiological reprogramming characterized by higher fat-free mass and lower basal metabolic rate.

These findings establish that early-life nutrition programs lifelong multisystem resilience and provide a mechanistic mandate for policies that limit added sugar exposure in the first 1,000 days—an intervention that extends well beyond preventing childhood obesity to shaping health across the life course.

## Data Availability

The deidentified participant data and data dictionaries used in this study are available from UK Biobank, HRS, and NHANES. Data are available upon publication, via formal application to the official websites: UK Biobank (https://www.ukbiobank.ac.uk/), HRS (https://hrs.isr.umich.edu/), NHANES (https://wwwn.cdc.gov/nchs/nhanes/). Access requires approval of a research proposal and compliance with each database's data access agreement. Additional documents such as the study protocol and statistical analysis plan are not available separately. Software code is available from the corresponding author upon reasonable request.

## Declarations

### Contributors

Yi. Z, D.C and X.Q were responsible for the concept and design of the study and were involved in writing the original draft of the manuscript. Yi. Z, X.C, and X.L conducted the statistical analysis, while D.C verified the data. X.Q obtained the funding and provided supervision throughout the research. All authors contributed to the acquisition, analysis, or interpretation of the data, as well as the review of the manuscript.

## Acknowledgments

This research has been conducted using the UK Biobank (Application Number 73201), HRS, and NHANES. We especially thank the all the participants and all the people involved in building these studies.

## Ethics approval and consent to participate

The UK Biobank was approved by the North West Center for Research Ethics Committee (11/NW/0382). The Health and Retirement Study (HRS) was approved by the University of Michigan Institutional Review Board (HUM00162208). The NHANES was approved by the National Center for Health Statistics Research Ethics Review Board. All participants signed an informed consent.

## References

1. Barker DJ. The origins of the developmental origins theory. J Intern Med. 2007;261(5):412–417. doi:10.1111/j.1365-2796.2007.01809.x

2. Gluckman PD, Hanson MA, Cooper C, Thornburg KL. Effect of in utero and early-life conditions on adult health and disease. N Engl J Med. 2008;359(1):61–73. doi:10.1056/NEJMra0708473

3. Guideline: Sugars Intake for Adults and Children. Geneva: World Health Organization; 2015.

4. DiNicolantonio JJ, O’Keefe JH, Lucan SC. Added sugar: the contributions of the food industry and the public health. N Engl J Med. 2015;372(16):1508–1512. doi:10.1056/NEJMp1501124

5. Gracner T, Boone C, Gertler PJ. Sugar rationing in early life and risk of type 2 diabetes and hypertension in adulthood: a natural experiment from post-war Britain. Science. 2024;384(6695):eadd5423. doi:10.1126/science.add5423

6. Zheng Y, Liu S, Wang L, et al. Early-life sugar restriction and long-term cardiovascular health: a natural experiment using UK sugar rationing. BMJ. 2025;388:e079832. doi:10.1136/bmj-2024-079832

7. Zheng Y, Liu S, Wang L, et al. Sugar rationing in the first 1000 days and risk of metabolic dysfunction-associated steatotic liver disease and cirrhosis: a natural experiment. Clin Gastroenterol Hepatol. 2025;23(4):612–621. doi:10.1016/j.cgh.2024.11.008

8. Chen H, Zhang J, Li W, et al. Early-life dietary sugar restriction and adult respiratory health: evidence from the UK sugar rationing natural experiment. Am J Clin Nutr. 2025;121(3):567–576. doi:10.1016/j.ajcnut.2024.12.015

9. Tang H, Zhang X, Huang J, et al. Sugar rationing during the first 1000 days of life and lifelong risk of heart failure. Nat Commun. 2026;17(1):1894. Published 2026 Jan 21. doi:10.1038/s41467-026-68713-9

10. Navratilova HF, Whetton AD, Geifman N. Dietary sugar exposure in early life and risk of adult mental health disorders: UK Biobank cohort study. medRxiv. Posted January 20, 2026. doi:10.64898/2026.01.20.26344391

11. Heijmans BT, Tobi EW, Stein AD, et al. Persistent epigenetic differences associated with prenatal exposure to famine in humans. Proc Natl Acad Sci USA. 2008;105(44):17046–17049. doi:10.1073/pnas.0806560105

12. You J, Guo Y, Zhang Y, et al. Plasma proteomic profiles predict individual future health risk. Nat Commun. 2023;14(1):7817. Published 2023 Nov 28. doi:10.1038/s41467-023-43575-7

13. Thompson DJ, Wells DK, Selzam S, et al. UK Biobank release and systematic evaluation of optimised polygenic risk scores for 53 diseases and quantitative traits. medRxiv. doi: 10.1101/2022.06.16.22276246

14. Moreno Villares JM. Los mil primeros días de vida y la prevención de la enfermedad en el adulto [Nutrition in early life and the programming of adult disease: the first 1000 days]. Nutr Hosp. 2016;33(Suppl 4):337. Published 2016 Jul 12. doi:10.20960/nh.337

15. Manning BD, Toker A. AKT/PKB Signaling: Navigating the Network. Cell. 2017;169(3):381–405. doi:10.1016/j.cell.2017.04.001

16. Beenken A, Mohammadi M. The FGF family: biology, pathophysiology and therapy. Nat Rev Drug Discov. 2009;8(3):235–253. doi:10.1038/nrd2792

17. Newgard CB. Interplay between lipids and branched-chain amino acids in development of insulin resistance. Cell Metab. 2012;15(5):606–614. doi:10.1016/j.cmet.2012.01.024

18. Wang TJ, Larson MG, Vasan RS, et al. Metabolite profiles and the risk of developing diabetes. Nat Med. 2011;17(4):448–453. doi:10.1038/nm.2307

19. Kensara OA, Wooton SA, Phillips DI, et al. Substrate-energy metabolism and metabolic risk factors for cardiovascular disease in relation to fetal growth and adult body composition. Am J Physiol Endocrinol Metab. 2006;291(2):E365–E371. doi:10.1152/ajpendo.00599.2005

20. Gallagher D, Heymsfield SB, Heo M, Jebb SA, Murgatroyd PR, Sakamoto Y. Healthy percentage body fat ranges: an approach for developing guidelines based on body mass index. Am J Clin Nutr. 2000;72(3):694–701. doi:10.1093/ajcn/72.3.694

